# The Challenges of Episodic Office-based Blood Pressure Measurement for the Management of Hypertension

**DOI:** 10.1101/2021.08.18.21262255

**Authors:** Yuan Lu, George C. Linderman, Shiwani Mahajan, Yuntian Liu, Bobak Mortazavi, Chenxi Huang, Rohan Khera, Erica S. Spatz, Harlan M. Krumholz

## Abstract

**Importance:** Clinicians use blood pressure (BP) readings obtained during clinical encounters to detect hypertension and determine the adequacy of treatment. Variations in office-based BP measurements may obscure a hypertension diagnosis or overwhelm a signal of treatment response.

**Objectives:** To quantify visit-to-visit variability (VVV) in BP values and its association with patient factors in real-world practice.

**Design, Setting and, Participants:** Retrospective cohort analysis of adult patients (age ≥18 years) with at least two outpatient visits in the Yale-New Haven Health System between January 1, 2014 to October 31, 2018.

**Main Outcome and Measures:** Patient-level measures of VVV included standard deviation (SD) and coefficient of variation (CV) of a given patient’s BP across visits. We introduced a metric to determine the VVV between any two visits (dyad) to characterize the BP information that clinicians have as they formulate their recommendations. Dyad-level measures of VVV included difference, absolute difference, standardized difference, and absolute standardized difference between the two visits of a dyad.

**Results:** The study population included 537,245 adults, with a total of 7,721,864 BP measurements. The mean age was 53.4 years (SD of 19.0), 60.4% were women, 69.4% were non-Hispanic White, and 18.1% with hypertension treatment. At the patient level, the mean intra-individual SD and CV were 10.6 mmHg and 0.08 mmHg. At the dyad level, the mean difference, absolute difference, standardized difference, and absolute standardized difference were -0.7 mmHg, 11.6 mmHg, 0 mmHg, and 0.09 mmHg, respectively. Given the observed VVV, if an antihypertensive medication truly reduced a patient’s SBP by 10 mmHg (the average BP-lowering effect reported in previous review), clinicians would expect to observe a reduction of SBP < 5mm Hg at the next visit 36.9% of the time. In the multivariable linear regression model, only 2% of the variance in absolute standardized difference was attributable to patient characteristics.

**Conclusions and Relevance:** The large VVV poses challenges for diagnosis, treatment, and monitoring of patients with hypertension based on BP readings in outpatient settings, supporting recent guidelines recommending home BP monitoring and ambulatory BP monitoring as out-of-office alternatives to establish diagnosis of hypertension and BP control.

**KEY POINTS:** *Question:* What is the visit-to-visit variability (VVV) in blood pressure (BP) values and its association with patient factors in real-world practice?

*Findings:* In this retrospective cohort analysis that included 537,245 adults and 7,721,864 office-based BP measurements from a large health system, marked VVV was observed in BP values and the median absolute change between two consecutive visits was about 12 mmHg. The VVV was not significantly associated with patient demographic and clinical characteristics.

*Meaning:* The large VVV poses challenges for diagnosis, treatment, and monitoring of patients with hypertension based on BP readings in outpatient settings, supporting recent guidelines recommending home BP monitoring and ambulatory BP monitoring as out-of-office alternatives to establish a new diagnosis of hypertension and BP control.

## BACKGROUND

In current practice, clinicians generally rely on blood pressure (BP) readings obtained during clinical encounters to detect elevated BP levels and the response to treatment strategies. The challenge is that office-based BP measurements vary in ways that can obscure a hypertension diagnosis or overwhelm a signal of treatment response.^1,2^ This variation can derive from biological fluctuations, and variation in timing, context, and method of measurement.^3,4^ This variation has led to the recommendation that BP changes be evaluated with home BP monitoring (HBPM) and ambulatory BP monitoring (ABPM) as out-of-office alternatives,^3,5^ but that is not common practice in part due to the cost and limited access to testing.^6^

Several studies, primarily using data collected in prospective studies, have documented visit-to-visit variability (VVV) in BP values and its association with the risk of adverse outcomes.^7-11^ For example, in the ALLHAT trial, the middle quintile of the systolic BP (SBP) standard deviation was approximately 10 mmHg and a higher VVV of SBP was associated with an increased risk of mortality.^11^ However, information is lacking on the individual BP measurement variation in real-world practice, where BP measurement, its timing, and context are less standardized. The information is important because, beyond its prognostic value, the real-world BP variation can represent noise that impairs clinical management.

Accordingly, we sought to describe real-world VVV using data from 537,245 outpatients with more than 7 million office-based BP measurements during a 5-years period from a large health system with a diverse patient population. Specifically, we quantified the VVV using different measures and investigated patient factors associated with VVV in real-world practice. We hypothesized that there is large VVV in office-based outpatient BP measurements and patient characteristics do not substantially explain variance in VVV.

## METHODS

### Study population and data source

The study population consisted of adult patients (age ≥18 years) with at least two outpatient visits in the Yale-New Haven Health System (YNHHS), from January 1, 2014 to December 31, 2018. YNHHS is Connecticut’s largest healthcare system, which provides care for approximately 3 million residents across Connecticut, New York, and Rhode Island. All YNHHS practice sites use Epic Corporations (Madison, WI) electronic health record system, so that every visit is integrated into the data repository queried for this study.

### Assessment of VVV of blood pressure

We assessed VVV of BP based on patient-level and dyad-level measures of variation. Consistent with previous studies of VVV, ^7-11^ patient-level measures of variation included standard deviation (SD) and coefficient of variation (CV) of a given patient’s BP across visits. SD was defined as the SD of all BP measurements for each patient, without restriction on the time duration between visits; CV was defined as SD divided by the mean of all BP measurements for each patient. If there were two or more BP measurements from the same visit, the mean BP was used as the value for that visit.

We further used measures of variation based on dyads formed by pairs of any two consecutive visits for the same patient occurring within a 90-day window. A patient might have multiple dyads of BP measurements during the study period. By using dyad-level measures, we quantified the variation that a clinician would observe when comparing the BP to the previous visit in diagnosis and management of hypertension by considering these dyads.

Dyad-level measures of variation included difference, absolute difference, standardized difference, absolute standardized difference, stage difference, and absolute stage difference. The difference was defined as the difference between BP measured at the first and second visits of a dyad; absolute difference was defined as the absolute value of difference. Standardized difference was defined as the difference between BP measured at the first and second visits of a dyad, divided by the BP at the first visit; absolute standardized difference was defined as the absolute value of standardized difference. Stage difference was defined as the number of hypertension stages changed between the first and second visits of a dyad; absolute stage difference was defined as the absolute value of the stage difference. Hypertension stages included normal BP (SBP <120 mmHg), elevated BP (SBP 120-129 mmHg), stage 1 hypertension (SBP 130-139 mmHg), stage 2 hypertension (SBP >=140 mmHg), and hypertension crisis (SBP >=180 mmHg). Detailed definitions of patient-level and dyad-level measures of variation are described in Supplemental Table S1. For the main analysis, we focused on SBP in calculating the dyad-level of measures for simplicity. We computed patient-level and dyad-level measures of variation as summaries (mean, median, interquartile range).

### Covariates

We computed VVV among subgroups formed by age, sex, race, ethnicity, treatment status, hypertensive stage, time between visits, and comorbidities (hypertension, hyperlipidemia, diabetes, coronary artery disease, and chronic kidney disease). The number of antihypertensive medications was used to determine treatment status. In particular, a dyad was labeled as “Increased treatment,” “Decreased treatment,” “Same treatment,” or “No treatment” depending on whether between the two visits, the number of antihypertensive medications increased, decreased, were both the same non-zero value, or were both zero, respectively. Comorbidities were identified using International Classification of Diseases, Ninth Revision or International Classification of Diseases, Tenth Revision codes listed in Supplemental Table S2. Comorbidities were defined as any diagnosis during the study period.

### Statistical analysis

We first describe the sociodemographic and clinical characteristics of the study population. We plotted the mean SBP and diastolic BP (DBP) distributions across each patient’s visits, time between visits, length of follow-up, and number of visits. We calculated various patient-level and dyad-level measures of variation, and then assessed how these measures varied by patient characteristics. Since patients might have different number of visits and dyads during the study period, we randomly selected a dyad for each patient when calculating the summary statistics of dyad-level measures of variation. We conducted two additional sensitivity analyses to test robustness of our results – the first used a different random dyad for each patient and the second used all dyads for each patient in calculating the dyad-level measures of variation.

To assess the extent to which VVV was explained by patient characteristics, we developed a multivariable linear regression model where we regressed the absolute standardized difference onto sex, age, race, treatment status, time from last visit, and comorbidities. The R^2^ was then reported as a metric of the variance in VVV that is explained by these covariates.

Previous systematic review of clinical trials reported that different antihypertensive mediations had an average effect of lowering SBP by approximately 10 mmHg.^12^ We estimated that if an antihypertensive medication truly reduced a patient’s SBP by 10 mmHg, the proportion of times that clinicians would expect to observe a SBP reduction of less than 5 mmHg (a change indicating no difference in BP had occurred) at the next visit given the observed VVV (**Supplemental Text**). We also estimated the proportion of times that clinicians would expect to observe no reduction in patients’ SBP at the next visit. Lastly, we estimated the number of visits it would take to be 80% certain that the true BP had decreased by 10 mmHg, assuming BP measurements in these visits were independent.

All analyses were conducted using R 4.0. All statistical testing was 2-sided at a significance level of 0.05. Institutional review board approval for this study was obtained through the Yale University Human Investigation Committee. The study followed the guidelines for cohort studies, described in the Strengthening the Reporting of Observational Studies in Epidemiology (STROBE) Statement: guidelines for reporting observational studies.^13^

## RESULTS

### Population characteristics

The study population included 537,245 adults (Supplemental Figure S1), with a total of 7,721,864 BP measurements over the study period. The mean age was 53.4 (SD 19.0) years, 60.4% were women, 69.4% were non-Hispanic White, and 18.1% received treatment for hypertension. Patients had a mean body mass index of 28.4 (5.9) kg/m^2^ and 22.6%, 8.0%, 9.7%, and 5.6% had a history of hypertension, diabetes, hyperlipidemia, and coronary artery disease, respectively (Table 1).

**Table 1.** Characteristics of study population at the patient-level.

| **Characteristics** | **Overall (N=537,245)** |
| --- | --- |
| **Age in years, mean (SD)** | 53.4 (19.0) |
| **Body mass index, mean (SD)** | 28.4 (5.9) |
| **Systolic BP across visits, mean (SD)** | 125.2 (12.8) |
| **Diastolic BP across visits, mean (SD)** | 75.1 (7.5) |
| **Number of visits per patient, mean (SD)** | 13.3 (15.6) |
| **Sex, N (%)** |  |
| Female | 324543 (60.4%) |
| Male | 212675 (39.6%) |
| Unknown | 27 (0.0%) |
| **Race/Ethnicity, N (%)** |  |
| Non-Hispanic Asian | 14880 (2.8%) |
| Non-Hispanic Black | 61968 (11.5%) |
| Non-Hispanic White | 373083 (69.4%) |
| Hispanic | 59797 (11.1%) |
| Other/Unknown | 27517 (5.1%) |
| **Insurance, N (%)** |  |
| Private | 294725 (54.9%) |
| Public | 216299 (40.3%) |
| Self-pay | 11765 (2.2%) |
| Other/Unknown | 14456 (2.7%) |
| **Comorbidities, N (%)** |  |
| Hypertension | 121490 (22.6%) |
| Hyperlipidemia | 52149 (9.7%) |
| Diabetes | 43065 (8.0%) |
| Chronic kidney disease | 9141 (1.7%) |
| Coronary artery disease | 30082 (5.6%) |
| Abbreviations: SD, standard deviation; BP, blood pressure. |  |

### Distribution of SBP measurements

Overall, the mean (SD) number of visits per patient was 13.3 (15.6), over an average period of 2.4 (1.6) years. The average of the mean SBPs and DBPs across each patient’s visits was 125 (12.8) mmHg and 75.1 (7.5) mmHg. The distributions of SBPs, time between visits, length of follow-up, and number of visits are shown in **Error! Reference source not found**.. The 20 most common primary diagnoses are listed in Supplemental Table S3, among which essential hypertension, hyperlipidemia, diabetes mellitus, cough, acute upper respiratory infection were the top five common diagnoses.

### VVV in SBP measurements

The distributions of patient-level and dyad-level measures of variation are shown in **Error! Reference source not found**.. Overall, the mean (SD) intra-individual SD and CV were 10.6 (5.1) mmHg and 0.08 (0.04) mmHg, and the median (interquartile range [IQR]) intra-individual SD and CV were 10.0 (7.3, 13.2) mmHg and 0.08 (0.06, 0.10) mmHg (Supplemental Table S4). These patient-level measures of variation did not vary substantially across patient subgroups (**Error! Reference source not found**.). For example, the intra-individual SD was 10.3 (4.9) mmHg among women and 10.9 (5.2) mmHg men, and the intra-individual CV was 0.08 (0.04) mmHg for both women and men.

At the dyad-level, the mean (SD) difference, absolute difference, standardized difference, and absolute standardized difference were -0.7 (15.6) mmHg, 11.6 (10.4) mmHg, 0 (0.1) mmHg, and 0.09 (0.08) mmHg, respectively, and the median (IQR) difference, absolute difference, standardized difference, and absolute standardized difference were 0 (−10, 8) mmHg, 10.0 (4, 17) mmHg, 0 (−0.08, 0.07) mmHg, and 0.07 (0.03, 0.13) mmHg, respectively (Figure 2 and Supplemental Table S5). These dyad-level measures of variation also did not vary substantially by patient characteristics and treatment status (**Error! Reference source not found**.). For example, the mean (SD) difference, absolute difference, standardized difference, and absolute standardized difference were -0.8 (18.0) mmHg, 13.6 (11.8) mmHg, 0 (0.1) mmHg, and 0.1 (0.09) mmHg among patients who were treated with the same antihypertensive medications during two visits and -0.4 (15.0) mmHg, 11.2 (10.0) mmHg, 0 (0.1) mmHg, 0.09 (0.08) mmHg among those who were untreated in the two visits. However, these measures differed by BP stage of previous visit. Among patients with mean SBP <119 mmHg, the mean (SD) difference, absolute difference, standardized difference, and absolute standardized difference were 0.6 (12.9) mmHg, 9.7 (8.6) mmHg, 0 (0.10) mmHg, and 0.08 (0.07) mmHg, whereas among those with mean SBP >180 mmHg these measures were -32.3 (23.5) mmHg, 33.9 (21.1) mmHg, -0. 17 (0.12) mmHg, and 0.18 (0.11) mmHg, respectively. The sensitivity analyses using a different random dyad or all dyads for each patient in calculating the dyad-level measures of variation showed consistent results as the main analysis (Supplemental Table S6-S7).

**Figure 1.**
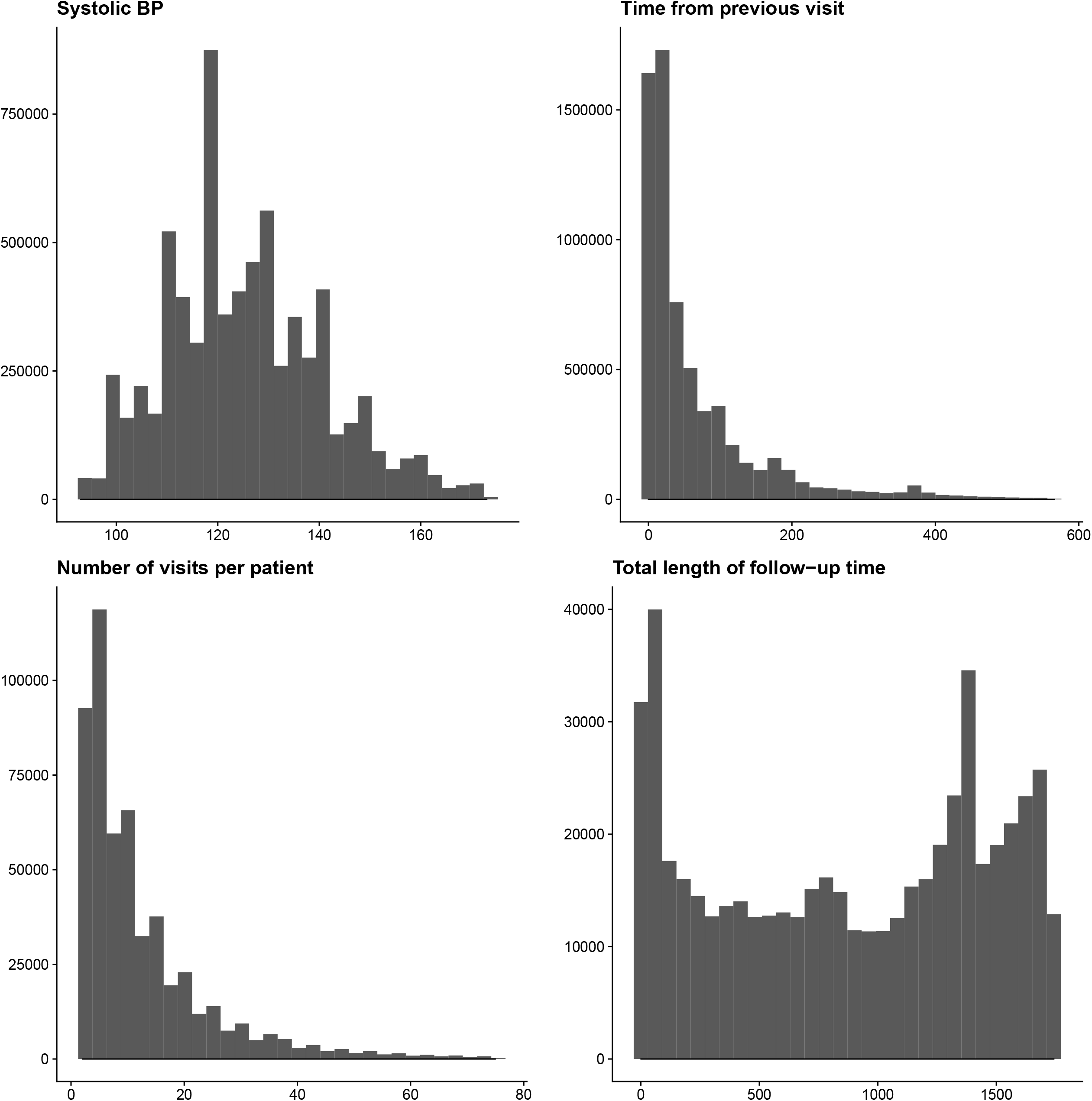
Distributions of systolic blood pressure (mmHg) across all visits, time (days) between visits, number of visits per patient, and total follow-up time (days) per patient.

**Figure 2.**
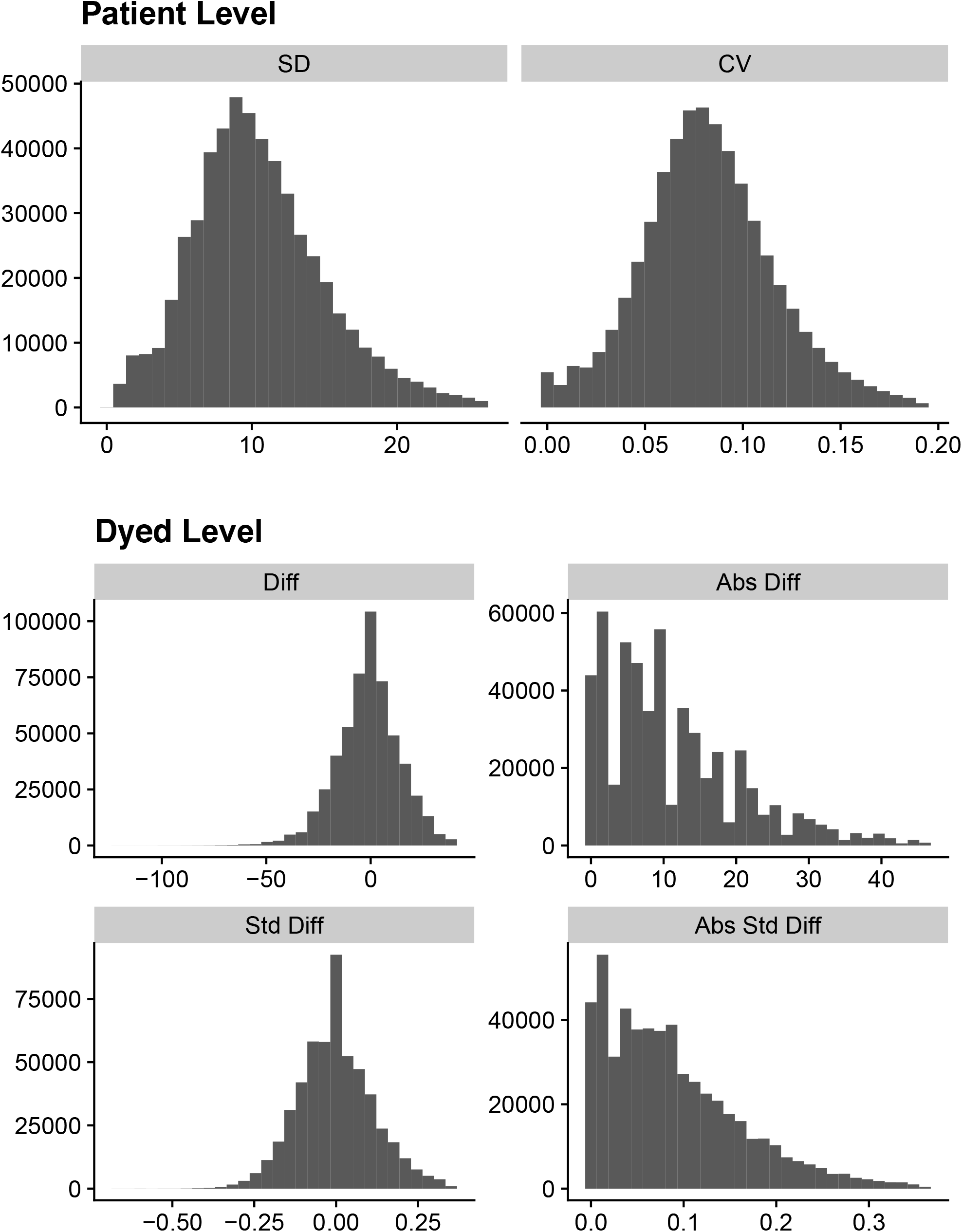
Distributions of patient-level and dyad-level measures of variation. Abbreviations: SD, Standard Deviation; CV, Coefficient of Variation; Diff, Difference; Abs Diff, Absolute Difference; Std Diff, Standardized Difference; Abs Std Diff, Absolute Standardized Difference.

**Figure 3.**
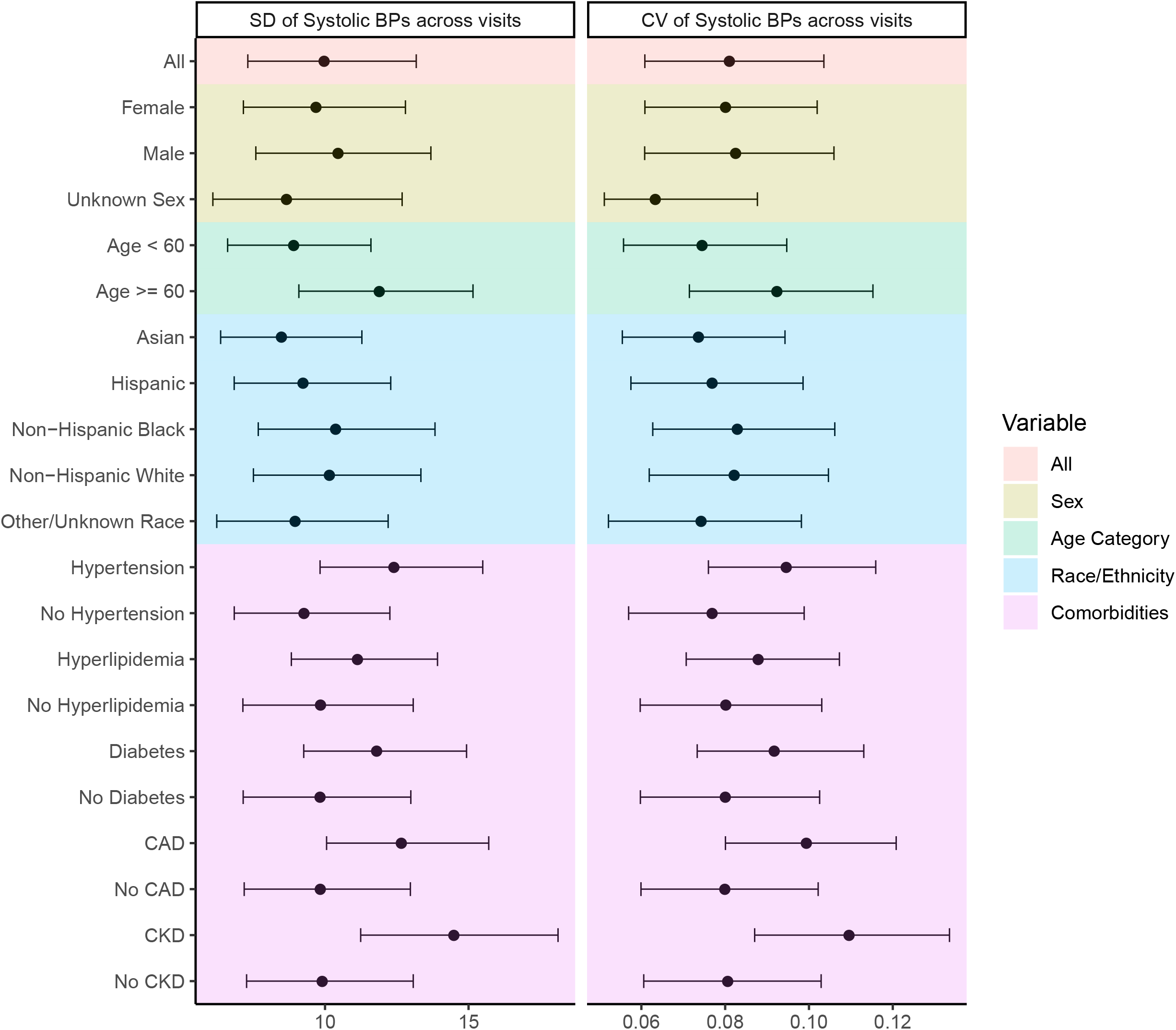
Patient-level measures of variation (median and interquartile range) across patient subgroups. Abbreviations: BP, blood pressure; CAD, coronary artery disease; CKD, chronic kidney disease; CV, coefficient of variation.

**Figure 4.**
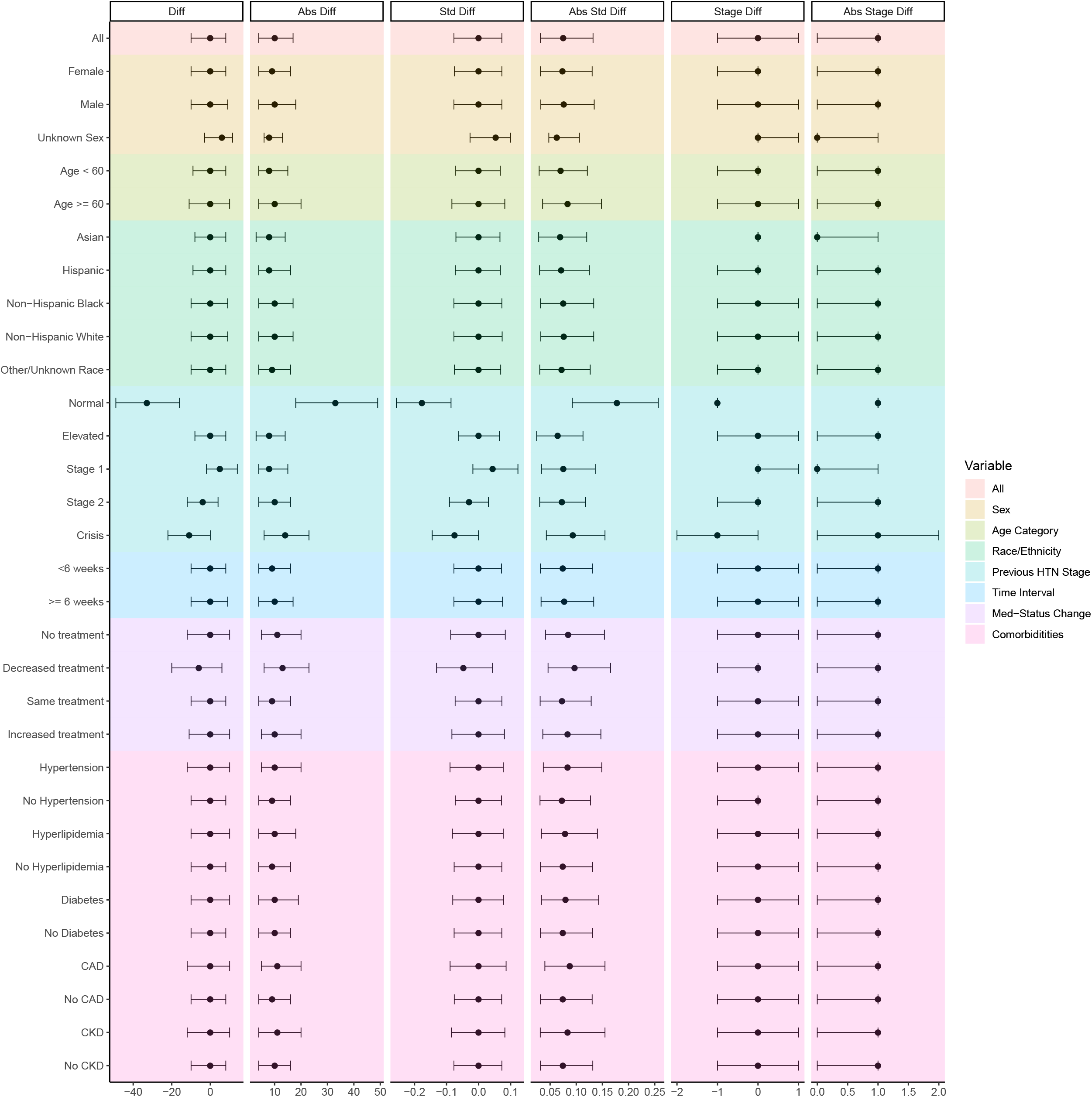
Dyad-level measures of variation (median and interquartile range) across patient subgroups. Abbreviations: Diff, Difference; Abs Diff, Absolute Difference; Std Diff, Standardized Difference; Abs Std Diff, Absolute Standardized Difference; Stage Diff, Stage Difference; Abs Stage Diff, Absolute Stage Difference; CAD, coronary artery disease; CKD, chronic kidney disease.

### Variance in VVV explained by covariates

The R^2^ for the multivariable linear regression model of absolute standardized difference onto covariates was 0.02, suggesting that 2% of the variance in this VVV was attributable to these measured covariates. The results did not change substantially by stage of hypertension.

If an antihypertensive medication reduced a patient’s SBP by 10 mmHg, given the observed VVV, clinicians would expect to observe no reduction in patient’s SBP at the follow-up visit 25.2% of the time. Clinicians would expect to observe a SBP reduction of < 5 mmHg at the follow-up visit 36.9% of the time. It would take approximate 4 separate visits to be 80% certain that the true BP had decreased by 10 mmHg, assuming BP measurements in these visits were independent.

## DISCUSSION

In this study of real-world outpatient BP VVV, we demonstrate marked variability that would largely overwhelm the detection of treatment effects between two visits. The median absolute change between two consecutive visits was about 12 mmHg, independent of treatment changes. Given that antihypertensive drugs tend to reduce BP less than this variation, patients and their physicians would routinely not be able to determine whether there is a treatment response based on measurements at a follow-up visit. Moreover, for patients in a decision-making zone of BP, the VVV was sufficiently large to have many patients pass in and out of treatment range. The VVV has received attention because of its prognostic importance, but it also has immense implications for hypertension treatment management.

This study extends the literature in several ways. We introduce the evaluation of VVV in consecutive visit dyads. The rationale is that clinicians commonly compare the current BP with the prior BP when evaluating an intervention, even as they may also look at them in the context of other prior measurements. Our analysis provides evidence of marked variability in real-world office-based BP measurements using large, contemporary data from a large health system. It also demonstrates more clearly the challenges VVV may pose for diagnosis, treatment, and monitoring of patients with hypertension based on BP readings in an outpatient setting. Multiple previous studies have demonstrated VVV, and its prognostic significance, using clinical trial data and data from longitudinal cohort studies.^7,9,11,14,15^ For example, by conducting secondary analysis of the data from the ALLHAT trial, Muntner et al showed large variation in the degree of VVV across all measurements among study participants and that higher VVV of SBP was associated with increased risk of cardiovascular disease and mortality.^11^

A notable finding of this study was that the mean VVV of real-world BP measurements was similar to that reported in the middle quintile of ALLHAT trial and the quintiles of VVV were similar in the two studies (8.7 to 11.0 mmHg in ALLHAT compared with 10.6 mmHg in our study). It might have been expected that VVV in the trial is less than what would have been observed in the real-world setting with standardization of measurement. Although it is challenging to perform a direct comparison between the studies, the fact that the VVV is largely similar suggests that greater standardization of clinician-measured BP may not decrease this variation further.

The cause of VVV is likely a result of many factors. Factors associated with arterial stiffness, including older age, female sex and higher pulse pressure, have been shown to be associated with VVV suggesting that alterations in vascular function may contribute to greater VVV.^10,15^ However, in this study, we show that much of the VVV in the real-world population is not related to patient factors, suggesting the need for a broad strategy to standardize BP measurements in every patient. VVV might also be the result of seasonal climatic changes, patient adherence and response to antihypertensive treatment in treated patients, or may reflect the inconsistencies in the timing, context, measurement device, and setting of BP measurement.^16^ The bottom line, though, is that unless we can reduce the noise, we are unlikely to be able to use episodic outpatient measurements to make clinical decisions and improve long-term BP stabilization.

The implications of the study are that it is difficult to detect treatment change in routine practice, which commonly involves having patients return for a BP check. The VVV can obscure meaningful changes following interventions. For example, if an antihypertensive medication might be expected to reduce SBP by 10 mmHg, a responder might be expected to have a reduction less than 5 mmHg in 37% of the time given the VVV. In fact, it would take approximate 4 separate visits to be 80% certain that the real BP had declined by 10 mmHg. As such, the common practice of episodic, office-based measurement may be highly insensitive in detecting the types of changes that might be expected with interventions.

The large VVV poses a great challenge for clinicians to determine hypertension status, therapy indications, and intervention responses in actual practice. This variation may be a key reason of why office-based management of hypertension is so difficult and why BP control is not more successful. Even though out-of-office HBPM and ABPM are now recommended,^3,5^ the vast majority of hypertension around the world is measured and managed in office-based settings. Understanding the VVV in real-world practice is a critical first step toward developing the necessary strategies to support decision-making amid an amplitude of change that may frequently cross BP categories and be larger than the expected effect of many interventions.

There are several options to address the problem. Current clinical decisions about titration of antihypertensive medications are often made on the basis of one or two office-based BP measurements. Using an average of several BP measurements may reduce within-patient variability and improve management decisions,^17^ although it may not always be possible. Artificial intelligence can be used to denoise the variation,^18^ but it still requires more BP measures. Having people return at same time of day and day of week and taking seasonal changes into account may also potentially reduce some variation. Finally, using ABPM that measures BP automatically and continually while patients perform daily activities could provide a better method for diagnosing hypertension and evaluating the impact of antihypertensive treatment.^5^ A recent systematic review has shown that ABPM has the highest sensitivity and specificity in detecting hypertension when comparing with office measurements of BP and HBPM.^1^ With the introduction of wearable cuffless BP measuring devices, such solutions may be on the horizon.^19^ When ABPM is not available, the American Heart Association and US Preventive Services Task Force recommend using HBPM in combination with office BP measurements as the preferred standard.^3,5^ The gap in using these methods lies in their implementation, which may require improvements in reimbursement and access to testing.^6^

### Limitations

This study has several potential limitations. First, the electronic health record data do not capture the exact device or technique used for BP measurement. However, our study focused on information that is available to the clinicians to make management decisions. Second, we did not consider the timing, context, or seasonal changes of BP measurements while estimating VVV. Third, treatment of hypertension was ascertained based on medication prescription data and we did not have data on patient adherence to medication. As previous studies reported that approximately 50% of the patients with hypertension do not take medications as prescribed,^20^ we might overestimate the true post-treatment VVV. Fourth, when calculating the patient- and dyad-level measures of VVV, patients who had few visits were weighted equally with the patients who had many visits.^21^ This may result in over or under-estimate of the true VVV in the study population. Finally, these data reflect the population catered by the YNHHS and may not necessarily be generalizable to other populations.

### Conclusion

There was marked VVV in real-world office-based BP measurements. This finding highlights the challenges VVV pose for diagnosis, treatment, and monitoring of patients with hypertension based on BP readings in outpatient settings and suggest the need to go beyond episodic clinic evaluation using HBPM and ABPM.

## Data Availability

Dr. Lu, Dr. Linderman, and Mr. Liu had full access to all the data in the study and take responsibility for the integrity of the data and the accuracy of the data analysis. Technical appendix, statistical code, and dataset available from the corresponding author.

## Access to Data Statement

Dr. Lu, Dr. Linderman, and Mr. Liu had full access to all the data in the study and take responsibility for the integrity of the data and the accuracy of the data analysis.

