## Supplemental material for "The Challenges of Episodic Office-based Blood Pressure Measurement for the Management of Hypertension"

Supplemental Text. Methods to estimate patients’ blood pressure change that clinicians would expect to observe during the follow-up visit

**Supplemental Figure S1.** Flow diagram of patient selection.

**Supplemental Table S1.** Definitions of patient-level and dyad-level measures of variation.

**Supplemental Table 2.** ICD-10 codes used to identify each comorbidity.

Supplemental Table S3. Top 15 most common primary diagnosis

Supplemental Table S4. Patient-level measures of variation across subgroups in the main analysis.

Supplemental Table S5. Dyad-level measures of variation across subgroups in the main analysis.

Supplemental Table S6. Dyad-level measures of variation across subgroups in the sensitivity analysis using a different random dyad for each patient.

Supplemental Table S7. Dyad-level measures of variation across subgroups in the sensitivity analysis using all dyads for each patient.

**Supplemental Appendix**

Supplemental Text. Methods to estimate patients’ blood pressure change that clinicians would expect to observe during the follow-up visit

Patient-level analysis:

Given the mean intra-individual standard deviation (SD) of systolic blood pressure (SBP) was 10.57 mmHg, we assumed the pre-treatment SBP followed a normal distribution of N (x1, 10.57), and post-treatment SBP followed a normal distribution of N (x2, 10.57). Then the SBP reduction after treatment would follow a normal distribution of N (x2-x1, 14.95). The SD of SBP reduction was calculated based on the pooled SD formula of (10.57^2+10.57^2) ^0.5 = 14.95.

If an antihypertensive treatment truly reduced the SBP by 10 mmHg, then x2-x1 = -10. The SBP reduction would follow a normal distribution of N (-10, 14.95). For a SBP change of -5 mmHg, we could then find out the corresponding percentile of 36.9%. That is, 36.9% of times that clinicians would observe an SBP change of -5 mmHg. Similarly, we found a SBP change of zero correspond to a percentile of 74.8%. That is, 25.2% of times that clinicians would observe no reduction in SBP.

Dyad-level analysis:

Similar approach can be applied to dyad level measure - consecutive difference. Instead of examining the variability across all measures for each patient, dyad-level focused on the two consecutive visits. We assumed the consecutive difference followed a normal distribution of N (0.66, 15.61). If an antihypertensive treatment truly reduced the SBP by 10 mmHg, then the SBP reduction under such treatment would follow a normal distribution N (-10+0.66 = -9.34, 15.61). For a SBP reduction of 5 mmHg, we could then find out the corresponding percentile of 42.2%. That is, 42.2% of times that clinicians would observe an SBP change of -5 mmHg. Similarly, we found a SBP change of zero correspond to a percentile of 66.4%. That is, 33.6% of times that clinicians would observe no reduction in SBP.

Supplemental Figure S1. Flow diagram of patient selection.

All adult patients seen outpatient at YNHHS during study period.

(n=859,716 patients)

Patients with all SBP measurements missing, below 40 mmHg, or above 300 mmHg.

(n=21,653)

Patients with at least two valid SBP measurements.

(n=838,063)

Patients with all visits separated by more than 90 days.

(n=300,818)

Patients with at least one valid dyad (consecutive SBP measurements within 90 days).

(n=537,245)

Supplemental Table S1. Definitions of patient-level and dyad-level measures of variation.

| **Measure** | **Definition** |
| --- | --- |
| **Patient-level measures of variation** | |
| Standard Deviation | The standard deviation of all BP measurements for each patient, without any restriction on the time duration between visits. |
| Coefficient of Variation | The standard deviation defined above, divided by the mean. |
| **Dyad-level measures of variation** | |
| Difference | The difference between systolic BP measured at the first and second visits of a dyad. |
| Absolute Difference | The absolute value of difference. |
| Standardized Difference | The difference between systolic BP measured at the first and second visits of a dyad, divided by the systolic BP at the first visit. |
| Absolute Standardized Difference | The absolute value of standardized difference. |
| Stage Difference | The number of hypertension stages changed between the first and second visits of a dyad. Specifically, we assign each patient an integer based on the systolic BP: 1, Normal, <119 mmHg; 2, Elevated, 120-129 mmHg; 3, Stage 1, 130-139 mmHg; 4, Stage 2,140-179 mmHg; 5, Crisis, >180 mmHg. For example, if the SBP was 135 on the first visit, and then 110 on the second, the stage changed from Stage 1 to Normal, so Stage Diff would be -2. |
| Absolute Stage Difference | The absolute value of stage difference |

Supplemental Table S2. ICD-10 codes used to identify each comorbidity.

|  | **ICD-9-CM** | **ICD-10-CM** |
| --- | --- | --- |
| Hypertension | 401, 402, 403, 404, 405, 642 | I10, I11, I12, I13, I15, I16, O13 |
| Diabetes mellitus | 249, 250 | E08, E09, E10 E11 E12 E13 |
| Hyperlipidemia | 272.0, 272.2, 272.3, 272.4 | E78.0, E78.2, E78.3, E78.4, E78.5 |
| Coronary artery disease | 414.0 | I25.1 |
| Chronic kidney disease | 585 | N18 |

Supplemental Table S3. Top 15 most common primary diagnosis

**Diagnosis Name**

Essential (primary) hypertension

Hyperlipidemia, unspecified

Diabetes mellitus

Cough

Acute upper respiratory infection, unspecified

Atrial fibrillation

Chronic ischemic heart disease without angina pectoris

Acute pharyngitis

Other malaise and fatigue

Disorder of urinary system, unspecified

Shortness of breath

Gastro-esophageal reflux disease without esophagitis

Obstructive sleep apnea (adult) (pediatric)

Gastro-esophageal reflux disease without esophagitis

Supplemental Table S4. Patient-level measures of variation across subgroups in the main analysis.

|  | **N** | **SD** | **CV** |
| --- | --- | --- | --- |
| **Total** | 537245 | 10.57 (5.05) | 0.08 (0.04) |
| **Sex** |  |  |  |
| Female | 324543 | 10.34 (4.94) | 0.08 (0.04) |
| Male | 212675 | 10.93 (5.21) | 0.08 (0.04) |
| **Age** |  |  |  |
| Age < 60 | 326826 | 9.39 (4.52) | 0.08 (0.03) |
| Age >= 60 | 210419 | 12.41 (5.29) | 0.09 (0.04) |
| **Race/Ethnicity** |  |  |  |
| Non-Hispanic Asian | 14880 | 9.12 (4.58) | 0.08 (0.03) |
| Non-Hispanic Black | 61968 | 11.09 (5.32) | 0.09 (0.04) |
| Non-Hispanic White | 373083 | 10.72 (5.01) | 0.08 (0.04) |
| Hispanic | 59797 | 9.90 (4.92) | 0.08 (0.04) |
| Other/Unknown Race | 27517 | 9.55 (5.15) | 0.08 (0.04) |
| **Hypertension status** |  |  |  |
| Hypertension | 121490 | 13.00 (4.83) | 0.10 (0.03) |
| No Hypertension | 415755 | 9.86 (4.89) | 0.08 (0.04) |
| **Hyperlipidemia status** |  |  |  |
| Hyperlipidemia | 52149 | 11.63 (4.23) | 0.09 (0.03) |
| No Hyperlipidemia | 485096 | 10.46 (5.12) | 0.08 (0.04) |
| **Diabetes status** |  |  |  |
| Diabetes | 43065 | 12.42 (4.71) | 0.09 (0.03) |
| No Diabetes | 494180 | 10.41 (5.05) | 0.08 (0.04) |
| **CAD status** |  |  |  |
| CAD | 30082 | 13.18 (4.70) | 0.10 (0.03) |
| No CAD | 507163 | 10.41 (5.03) | 0.08 (0.04) |
| **CKD status** |  |  |  |
| CKD | 9141 | 14.78 (6.13) | 0.11 (0.04) |
| No CKD | 528104 | 10.50 (5.00) | 0.08 (0.04) |

Abbreviations: SD, Standard Deviation; N, number; CV, Coefficient of Variation; CAD, coronary artery disease; CKD, chronic kidney disease.

Supplemental Table S5. Dyad-level measures of variation across subgroups in the main analysis.

|  | **Number** | **Diff** | **Abs Diff** | **Std Diff** | **Abs Std Diff** | **Stage Diff** | **Abs Stage Diff** |
| --- | --- | --- | --- | --- | --- | --- | --- |
| **Total** | 537245 | -0.66 (15.61) | 11.64 (10.42) | 0.00 (0.12) | 0.09 (0.08) | -0.04 (1.11) | 0.75 (0.82) |
| **Sex** |  |  |  |  |  |  |  |
| Female | 324543 | -0.59 (15.30) | 11.36 (10.27) | 0.00 (0.12) | 0.09 (0.08) | -0.03 (1.07) | 0.70 (0.81) |
| Male | 212675 | -0.77 (16.07) | 12.08 (10.62) | 0.00 (0.13) | 0.09 (0.08) | -0.05 (1.17) | 0.81 (0.83) |
| Unknown | 27 | 4.41 (10.87) | 9.59 (6.54) | 0.04 (0.09) | 0.08 (0.06) | 0.37 (0.74) | 0.44 (0.70) |
| **Age** |  |  |  |  |  |  |  |
| Age < 60 | 322433 | -0.59 (13.83) | 10.36 (9.17) | 0.00 (0.11) | 0.08 (0.07) | -0.04 (1.02) | 0.66 (0.78) |
| Age >= 60 | 214812 | -0.76 (17.96) | 13.57 (11.79) | 0.00 (0.14) | 0.10 (0.09) | -0.04 (1.23) | 0.87 (0.87) |
| **Race/Ethnicity** |  |  |  |  |  |  |  |
| Non-Hispanic Asian | 14880 | -0.54 (13.55) | 10.06 (9.10) | 0.00 (0.11) | 0.08 (0.07) | -0.04 (0.94) | 0.56 (0.75) |
| Non-Hispanic Black | 61968 | -0.69 (16.19) | 11.99 (10.90) | 0.00 (0.12) | 0.09 (0.08) | -0.04 (1.12) | 0.76 (0.82) |
| Non-Hispanic White | 373083 | -0.64 (15.81) | 11.83 (10.50) | 0.00 (0.12) | 0.09 (0.08) | -0.04 (1.13) | 0.77 (0.83) |
| Hispanic | 59797 | -0.70 (14.64) | 10.85 (9.85) | 0.00 (0.12) | 0.09 (0.08) | -0.04 (1.04) | 0.68 (0.79) |
| Other/Unknown Race | 27517 | -0.80 (14.61) | 10.91 (9.74) | 0.00 (0.12) | 0.09 (0.08) | -0.05 (1.05) | 0.69 (0.79) |
| **Hypertension stage** |  |  |  |  |  |  |  |
| Normal | 191949 | 6.28 (12.81) | 10.55 (9.61) | 0.06 (0.12) | 0.10 (0.09) | 0.55 (0.84) | 0.55 (0.84) |
| Elevated | 138799 | 0.56 (12.94) | 9.67 (8.62) | 0.00 (0.10) | 0.08 (0.07) | 0.10 (1.00) | 0.77 (0.65) |
| Stage 1 | 99683 | -3.37 (13.88) | 11.01 (9.10) | -0.03 (0.10) | 0.08 (0.07) | -0.43 (1.05) | 0.90 (0.69) |
| Stage 2 | 102740 | -11.38 (16.70) | 16.10 (12.20) | -0.07 (0.11) | 0.11 (0.08) | -0.89 (1.04) | 0.93 (1.01) |
| Crisis | 4074 | -32.26 (23.47) | 33.87 (21.07) | -0.17 (0.12) | 0.18 (0.11) | -1.20 (0.94) | 1.20 (0.94) |
| **Time between two visits** |  |  |  |  |  |  |  |
| <6 weeks | 361496 | -0.72 (15.58) | 11.59 (10.44) | 0.00 (0.12) | 0.09 (0.08) | -0.04 (1.10) | 0.73 (0.82) |
| >= 6 weeks | 175749 | -0.53 (15.66) | 11.75 (10.36) | 0.00 (0.12) | 0.09 (0.08) | -0.03 (1.13) | 0.77 (0.83) |
| **Treatment change between two visits** |  |  |  |  |  |  |  |
| No treatment | 461359 | -0.39 (15.01) | 11.20 (9.99) | 0.00 (0.12) | 0.09 (0.08) | -0.03 (1.09) | 0.72 (0.81) |
| Decreased treatment | 9471 | -0.76 (18.89) | 14.31 (12.36) | 0.01 (0.15) | 0.11 (0.10) | -0.03 (1.25) | 0.89 (0.87) |
| Same treatment | 47712 | -0.76 (17.99) | 13.59 (11.81) | 0.00 (0.14) | 0.10 (0.09) | -0.04 (1.23) | 0.87 (0.87) |
| Increased treatment | 18703 | -6.83 (19.95) | 16.28 (13.40) | -0.04 (0.14) | 0.11 (0.09) | -0.36 (1.24) | 0.92 (0.91) |
| **Hypertension status** |  |  |  |  |  |  |  |
| Hypertension | 121490 | -1.44 (18.42) | 13.98 (12.09) | -0.00 (0.14) | 0.10 (0.09) | -0.08 (1.25) | 0.90 (0.87) |
| No Hypertension | 415755 | -0.43 (14.68) | 10.96 (9.77) | 0.00 (0.12) | 0.09 (0.08) | -0.03 (1.07) | 0.70 (0.80) |
| **Hyperlipidemia status** |  |  |  |  |  |  |  |
| Hyperlipidemia | 52149 | -0.82 (16.62) | 12.58 (10.89) | 0.00 (0.13) | 0.10 (0.08) | -0.05 (1.20) | 0.86 (0.84) |
| No Hyperlipidemia | 485096 | -0.64 (15.50) | 11.54 (10.36) | 0.00 (0.12) | 0.09 (0.08) | -0.04 (1.10) | 0.73 (0.82) |
| **Diabetes status** |  |  |  |  |  |  |  |
| Diabetes | 43065 | -0.65 (17.53) | 13.13 (11.63) | 0.00 (0.13) | 0.10 (0.09) | -0.04 (1.21) | 0.86 (0.85) |
| No Diabetes | 494180 | -0.66 (15.43) | 11.51 (10.29) | 0.00 (0.12) | 0.09 (0.08) | -0.04 (1.10) | 0.74 (0.82) |
| **CAD status** |  |  |  |  |  |  |  |
| CAD | 30082 | -0.69 (18.49) | 14.08 (12.00) | 0.01 (0.14) | 0.11 (0.09) | -0.04 (1.27) | 0.90 (0.89) |
| No CAD | 507163 | -0.66 (15.42) | 11.50 (10.30) | 0.00 (0.12) | 0.09 (0.08) | -0.04 (1.10) | 0.74 (0.82) |
| **CKD status** |  |  |  |  |  |  |  |
| CKD | 9141 | -0.59 (19.54) | 14.27 (13.36) | 0.01 (0.15) | 0.11 (0.10) | -0.02 (1.21) | 0.81 (0.90) |
| No CKD | 528104 | -0.66 (15.53) | 11.60 (10.35) | 0.00 (0.12) | 0.09 (0.08) | -0.04 (1.11) | 0.75 (0.82) |

Abbreviations: Diff, Difference; Abs Diff, Absolute Difference; Abs Std Diff, Absolute Standardized Difference. CAD, coronary artery disease; CKD, chronic kidney disease.

Supplemental Table S6. Dyad-level measures of variation across subgroups in the sensitivity analysis using a different random dyad for each patient.

|  | **Number** | **Diff** | **Abs Diff** | **Std Diff** | **Abs Std Diff** | **Stage Diff** | **Abs Stage Diff** |
| --- | --- | --- | --- | --- | --- | --- | --- |
| **Total** | 537245 | -0.64 (15.61) | 11.65 (10.41) | 0.00 (0.12) | 0.09 (0.08) | -0.04 (1.11) | 0.75 (0.82) |
| **Sex** |  |  |  |  |  |  |  |
| Female | 324543 | -0.56 (15.31) | 11.38 (10.26) | 0.00 (0.12) | 0.09 (0.08) | -0.04 (1.07) | 0.70 (0.81) |
| Male | 212675 | -0.75 (16.05) | 12.06 (10.62) | 0.00 (0.12) | 0.09 (0.08) | -0.05 (1.17) | 0.81 (0.84) |
| Unknown | 27 | 5.67 (11.13) | 9.81 (7.56) | 0.05 (0.08) | 0.08 (0.06) | 0.41 (0.69) | 0.48 (0.64) |
| **Age** |  |  |  |  |  |  |  |
| Age < 60 | 322390 | -0.58 (13.86) | 10.39 (9.19) | 0.00 (0.11) | 0.08 (0.07) | -0.04 (1.02) | 0.67 (0.78) |
| Age >= 60 | 214855 | -0.73 (17.92) | 13.54 (11.76) | 0.00 (0.14) | 0.10 (0.09) | -0.04 (1.23) | 0.87 (0.87) |
| **Race/Ethnicity** |  |  |  |  |  |  |  |
| Non-Hispanic Asian | 14880 | -0.51 (13.63) | 10.14 (9.12) | 0.00 (0.11) | 0.09 (0.07) | -0.04 (0.93) | 0.55 (0.75) |
| Non-Hispanic Black | 61968 | -0.58 (16.14) | 11.95 (10.86) | 0.00 (0.12) | 0.09 (0.08) | -0.03 (1.12) | 0.76 (0.82) |
| Non-Hispanic White | 373083 | -0.64 (15.80) | 11.83 (10.49) | 0.00 (0.12) | 0.09 (0.08) | -0.04 (1.13) | 0.77 (0.83) |
| Hispanic | 59797 | -0.64 (14.70) | 10.90 (9.89) | 0.00 (0.12) | 0.09 (0.08) | -0.04 (1.04) | 0.68 (0.79) |
| Other/Unknown Race | 27517 | -0.81 (14.70) | 10.95 (9.83) | 0.00 (0.12) | 0.09 (0.08) | -0.05 (1.05) | 0.69 (0.80) |
| **Hypertension stage** |  |  |  |  |  |  |  |
| Normal | 191767 | 6.33 (12.85) | 10.60 (9.63) | 0.06 (0.12) | 0.10 (0.09) | 0.55 (0.84) | 0.55 (0.84) |
| Elevated | 139236 | 0.53 (12.94) | 9.67 (8.61) | 0.00 (0.10) | 0.08 (0.07) | 0.09 (1.00) | 0.77 (0.65) |
| Stage 1 | 99785 | -3.40 (13.88) | 11.00 (9.11) | -0.03 (0.10) | 0.08 (0.07) | -0.44 (1.05) | 0.90 (0.69) |
| Stage 2 | 102405 | -11.31 (16.64) | 16.04 (12.14) | -0.07 (0.11) | 0.11 (0.08) | -0.89 (1.04) | 0.93 (1.00) |
| Crisis | 4052 | -32.61 (23.81) | 34.09 (21.64) | -0.17 (0.12) | 0.18 (0.11) | -1.22 (0.97) | 1.22 (0.97) |
| **Time between two visits** |  |  |  |  |  |  |  |
| <6 weeks | 361983 | -0.70 (15.59) | 11.60 (10.44) | 0.00 (0.12) | 0.09 (0.08) | -0.04 (1.10) | 0.74 (0.82) |
| >= 6 weeks | 175262 | -0.51 (15.65) | 11.76 (10.34) | 0.00 (0.12) | 0.09 (0.08) | -0.03 (1.13) | 0.77 (0.83) |
| **Treatment change between two visits** |  |  |  |  |  |  |  |
| No treatment | 461691 | -0.39 (15.01) | 11.21 (9.99) | 0.00 (0.12) | 0.09 (0.08) | -0.03 (1.09) | 0.73 (0.81) |
| Decreased treatment | 9376 | -0.90 (18.81) | 14.34 (12.21) | 0.00 (0.14) | 0.11 (0.09) | -0.03 (1.25) | 0.89 (0.88) |
| Same treatment | 47496 | -0.68 (18.02) | 13.60 (11.84) | 0.00 (0.14) | 0.10 (0.09) | -0.05 (1.23) | 0.87 (0.87) |
| Increased treatment | 18682 | -6.52 (19.97) | 16.16 (13.42) | -0.03 (0.14) | 0.11 (0.09) | -0.34 (1.24) | 0.91 (0.90) |
| **Hypertension status** |  |  |  |  |  |  |  |
| Hypertension | 121490 | -1.40 (18.39) | 13.95 (12.07) | -0.00 (0.14) | 0.10 (0.09) | -0.08 (1.25) | 0.90 (0.87) |
| No Hypertension | 415755 | -0.41 (14.69) | 10.98 (9.77) | 0.00 (0.12) | 0.09 (0.08) | -0.03 (1.07) | 0.70 (0.80) |
| **Hyperlipidemia status** |  |  |  |  |  |  |  |
| Hyperlipidemia | 52149 | -0.78 (16.67) | 12.59 (10.94) | 0.00 (0.13) | 0.10 (0.08) | -0.05 (1.21) | 0.87 (0.85) |
| No Hyperlipidemia | 485096 | -0.62 (15.49) | 11.55 (10.34) | 0.00 (0.12) | 0.09 (0.08) | -0.04 (1.10) | 0.74 (0.82) |
| **Diabetes status** |  |  |  |  |  |  |  |
| Diabetes | 43065 | -0.44 (17.41) | 13.05 (11.53) | 0.01 (0.13) | 0.10 (0.09) | -0.03 (1.21) | 0.86 (0.86) |
| No Diabetes | 494180 | -0.65 (15.44) | 11.53 (10.29) | 0.00 (0.12) | 0.09 (0.08) | -0.04 (1.10) | 0.74 (0.82) |
| **CAD status** |  |  |  |  |  |  |  |
| CAD | 30082 | -0.59 (18.60) | 14.16 (12.08) | 0.01 (0.14) | 0.11 (0.09) | -0.03 (1.27) | 0.91 (0.89) |
| No CAD | 507163 | -0.64 (15.41) | 11.50 (10.28) | 0.00 (0.12) | 0.09 (0.08) | -0.04 (1.10) | 0.74 (0.82) |
| **CKD status** |  |  |  |  |  |  |  |
| CKD | 9141 | -0.43 (19.69) | 14.38 (13.46) | 0.01 (0.15) | 0.11 (0.10) | -0.01 (1.24) | 0.84 (0.91) |
| No CKD | 528104 | -0.64 (15.53) | 11.60 (10.34) | 0.00 (0.12) | 0.09 (0.08) | -0.04 (1.11) | 0.75 (0.82) |

Abbreviations: Diff, Difference; Abs Diff, Absolute Difference; Abs Std Diff, Absolute Standardized Difference. CAD, coronary artery disease; CKD, chronic kidney disease.

Supplemental Table S7. Dyad-level measures of variation across subgroups in the sensitivity analysis using all dyads for each patient.

|  | **Number** | **Diff** | **Abs Diff** | **Std Diff** | **Abs Std Diff** | **Stage Diff** | **Abs Stage Diff** |
| --- | --- | --- | --- | --- | --- | --- | --- |
| **Total** | 5004265 | -0.29 (16.15) | 12.11 (10.69) | 0.01 (0.13) | 0.10 (0.08) | -0.02 (1.14) | 0.77 (0.84) |
| **Sex** |  |  |  |  |  |  |  |
| Female | 3090062 | -0.26 (15.77) | 11.76 (10.51) | 0.01 (0.13) | 0.09 (0.08) | -0.02 (1.10) | 0.73 (0.83) |
| Male | 1914061 | -0.33 (16.74) | 12.67 (10.95) | 0.01 (0.13) | 0.10 (0.09) | -0.02 (1.19) | 0.83 (0.85) |
| Unknown | 142 | 1.57 (12.56) | 9.95 (7.78) | 0.02 (0.10) | 0.08 (0.06) | 0.11 (0.84) | 0.49 (0.69) |
| **Age** |  |  |  |  |  |  |  |
| Age < 60 | 2481740 | -0.26 (14.15) | 10.59 (9.39) | 0.00 (0.12) | 0.09 (0.08) | -0.02 (1.03) | 0.67 (0.78) |
| Age >= 60 | 2522525 | -0.31 (17.90) | 13.60 (11.64) | 0.01 (0.14) | 0.11 (0.09) | -0.02 (1.23) | 0.87 (0.88) |
| **Race/Ethnicity** |  |  |  |  |  |  |  |
| Non-Hispanic Asian | 109368 | -0.29 (14.06) | 10.40 (9.46) | 0.00 (0.12) | 0.09 (0.08) | -0.02 (0.95) | 0.56 (0.76) |
| Non-Hispanic Black | 716615 | -0.29 (16.73) | 12.48 (11.14) | 0.01 (0.13) | 0.10 (0.09) | -0.02 (1.15) | 0.78 (0.84) |
| Non-Hispanic White | 3453931 | -0.29 (16.27) | 12.24 (10.72) | 0.01 (0.13) | 0.10 (0.09) | -0.02 (1.15) | 0.79 (0.84) |
| Hispanic | 553312 | -0.28 (15.27) | 11.34 (10.23) | 0.01 (0.12) | 0.09 (0.08) | -0.02 (1.07) | 0.70 (0.81) |
| Other/Unknown Race | 171039 | -0.37 (15.14) | 11.34 (10.04) | 0.00 (0.12) | 0.09 (0.08) | -0.02 (1.08) | 0.70 (0.81) |
| **Hypertension stage** |  |  |  |  |  |  |  |
| Normal | 1786904 | 6.80 (13.37) | 11.11 (10.08) | 0.07 (0.13) | 0.10 (0.10) | 0.58 (0.87) | 0.58 (0.87) |
| Elevated | 1245612 | 1.11 (13.54) | 10.20 (8.97) | 0.01 (0.11) | 0.08 (0.07) | 0.15 (1.03) | 0.80 (0.66) |
| Stage 1 | 948820 | -2.97 (14.39) | 11.35 (9.32) | -0.02 (0.11) | 0.09 (0.07) | -0.40 (1.07) | 0.91 (0.69) |
| Stage 2 | 987717 | -11.14 (17.08) | 16.24 (12.34) | -0.07 (0.11) | 0.11 (0.08) | -0.89 (1.05) | 0.92 (1.01) |
| Crisis | 35212 | -32.72 (23.83) | 34.36 (21.39) | -0.17 (0.12) | 0.18 (0.11) | -1.23 (0.97) | 1.23 (0.97) |
| **Time between two visits** |  |  |  |  |  |  |  |
| <6 weeks | 3885610 | -0.34 (16.10) | 12.05 (10.68) | 0.01 (0.13) | 0.10 (0.08) | -0.02 (1.13) | 0.76 (0.84) |
| >= 6 weeks | 1118655 | -0.12 (16.31) | 12.28 (10.73) | 0.01 (0.13) | 0.10 (0.08) | -0.01 (1.17) | 0.81 (0.84) |
| **Treatment change between two visits** |  |  |  |  |  |  |  |
| No treatment | 3942930 | -0.12 (15.46) | 11.58 (10.23) | 0.01 (0.12) | 0.09 (0.08) | -0.01 (1.11) | 0.74 (0.82) |
| Decreased treatment | 113645 | -0.28 (19.21) | 14.56 (12.54) | 0.01 (0.15) | 0.11 (0.10) | -0.02 (1.26) | 0.90 (0.89) |
| Same treatment | 791254 | -0.27 (17.97) | 13.64 (11.70) | 0.01 (0.14) | 0.11 (0.09) | -0.02 (1.22) | 0.86 (0.87) |
| Increased treatment | 156436 | -4.59 (20.00) | 15.72 (13.19) | -0.02 (0.15) | 0.11 (0.09) | -0.25 (1.25) | 0.89 (0.90) |
| **Hypertension status** |  |  |  |  |  |  |  |
| Hypertension | 1786459 | -0.47 (18.23) | 13.82 (11.91) | 0.01 (0.14) | 0.11 (0.09) | -0.03 (1.25) | 0.89 (0.87) |
| No Hypertension | 3217806 | -0.19 (14.86) | 11.16 (9.82) | 0.01 (0.12) | 0.09 (0.08) | -0.01 (1.07) | 0.70 (0.81) |
| **Hyperlipidemia status** |  |  |  |  |  |  |  |
| Hyperlipidemia | 745767 | -0.32 (17.23) | 13.05 (11.25) | 0.01 (0.13) | 0.10 (0.09) | -0.02 (1.22) | 0.87 (0.86) |
| No Hyperlipidemia | 4258498 | -0.28 (15.95) | 11.94 (10.58) | 0.01 (0.13) | 0.10 (0.08) | -0.02 (1.12) | 0.75 (0.83) |
| **Diabetes status** |  |  |  |  |  |  |  |
| Diabetes | 786007 | -0.25 (18.04) | 13.60 (11.85) | 0.01 (0.14) | 0.10 (0.09) | -0.01 (1.23) | 0.87 (0.87) |
| No Diabetes | 4218258 | -0.30 (15.77) | 11.83 (10.44) | 0.01 (0.13) | 0.09 (0.08) | -0.02 (1.12) | 0.75 (0.83) |
| **CAD status** |  |  |  |  |  |  |  |
| CAD | 513488 | -0.25 (18.49) | 14.08 (11.99) | 0.01 (0.15) | 0.11 (0.09) | -0.01 (1.26) | 0.89 (0.89) |
| No CAD | 4490777 | -0.29 (15.86) | 11.88 (10.51) | 0.01 (0.13) | 0.09 (0.08) | -0.02 (1.12) | 0.75 (0.83) |
| **CKD status** |  |  |  |  |  |  |  |
| CKD | 245322 | -0.17 (19.71) | 14.88 (12.93) | 0.01 (0.15) | 0.11 (0.10) | -0.01 (1.25) | 0.87 (0.90) |
| No CKD | 4758943 | -0.29 (15.94) | 11.96 (10.54) | 0.01 (0.13) | 0.10 (0.08) | -0.02 (1.13) | 0.76 (0.83) |

Abbreviations: Diff, Difference; Abs Diff, Absolute Difference; Abs Std Diff, Absolute Standardized Difference. CAD, coronary artery disease; CKD, chronic kidney disease.
